# Predicting cancer patient mortality within 30 days of radiotherapy consultation to inform palliative radiotherapy fractionation decisions

**DOI:** 10.1101/2024.09.13.24313658

**Authors:** Kendall J. Kiser, Ashish Vaidyanathan, Matthew J. Schuelke, Joshua Denzer, Trudy Landreth, Christopher D. Abraham, Adam B. Wilcox

**Author notes:** **Corresponding Author** Adam Wilcox PhD. **Funding Statement** Support for this work was provided in part by a National Institutes of Health grant (NIH Research Portfolio Online Reporting Tools project number 75N95021D00028-0-759502200002-1). **Data Sharing Statement** This project involved secondary analysis of electronic health records. No data were collected as primary research data. To maintain patient record confidentiality, no data will be made publicly available.

## Abstract

**Background:** Radiation and medical oncologists evaluate patients’ risk of imminent mortality with scales like Karnofsky Performance Status (KPS) and predicate treatment decisions on these evaluations. However, we hypothesized that statistical models derived from structured electronic health record (EHR) data could predict patient deaths within 30 days of radiotherapy consultation better than models developed only with patient age and physician-reported KPS.

**Methods:** Clinical data from patients who consulted in a radiotherapy department from June 2018 – February 2024 were abstracted from EHR databases, including patient demographics, laboratory results, medications, comorbidities, KPS, cancer stages, oncologic treatment histories, oncologist notes, radiologist reports, and pathologist narratives. A subset of structured features known or believed to be associated with mortality were curated and used to train and test logistic regression, random forest, and gradient-boosted decision classifiers.

**Results:** Of 38,262 patients, 951 (2.5%) died within 30 days of radiotherapy consultation. From 34.5 gigabytes of tabular data, 2,977 clinical features were chosen or derived by a radiation oncologist, then reduced to 1,000 features using ANOVA F values. Using an event probability classification threshold of 0.2, optimized logistic regression, random forest, and gradient-boosted decision classifiers tested with high accuracy (0.97, 0.98, and 0.98, respectively) and F1 scores (0.50, 0.54, and 0.52). The areas under receiver operating and precision-recall curves for the random forest model were respectively 0.94 and 0.55, which outperformed a model trained only with patient age and KPS (0.61 and 0.06). Models prominently weighed features that were rationally associated with mortality.

**Conclusion:** Statistical models developed from a physician-curated feature space of structured EHR data predicted patient deaths within 30 days of radiotherapy consultation better than a model developed only with a patient’s age and physician-assessed KPS. With clinically explicable feature weights, these models could influence treatment decisions such as the length of palliative radiotherapy courses.

## Introduction

In some cancer patients, the risk of 30-day mortality is high. According to one meta-analysis of 88,000 patients, the risk in advanced cancer patients may be as high as 16%.^1^ Nevertheless, this estimate varies substantially with such factors as primary cancer site and patient performance status. Correctly identifying which cancer patients are at imminent mortality risk is difficult, and physicians are poor predictors.^2,3^

If it were possible to accurately estimate cancer patients’ 30-day mortality risk, this could meaningfully better their treatment by involving early palliative care, which has been demonstrated to improve quality of life and lengthen survival.^4-6^ It may also curtail unnecessary treatment. For example, radiation oncologists must decide whether to prescribe short or prolonged radiation therapy (RT) treatment courses to palliate cancer-related pain. Nearly 70% of patients treated with palliative RT report less pain, and RT prescribed in a single treatment is as likely to achieve pain relief as five or ten treatments (fractions),^7-10^ but the durability of relief is shorter^7,8,10^ and the risk of long-term complications (e.g. local tumor progression^11^) greater with a single treatment than a prolonged course. Complicating a radiation oncologist’s decision further, dose-intensified stereotactic ablative radiotherapy (SABR) provided better pain relief than conventionally-dose fractionation schedules in some trials,^12,13^ yet SABR planning requires more work is more costly. Patients who pass away during or immediately after treatment with prolonged or dose-intensified palliative courses may have been better served by a single fraction.^14^ One Surveillance, Epidemiology, and End Results (SEER) database study reported that 48% of patients who received palliative RT before enrolling in hospice received an RT course that was longer than the number of days they survived in hospice.^15^

Emerging evidence suggests that artificial intelligence (AI) may be able to predict patients’ 30-day mortality risk. For example, the University of Pennsylvania Health System trained an AI to predict patients’ six-month mortality risk with data from medical oncology patients’ electronic health records (EHR)^16^ and validated the AI’s efficacy at increasing end-of-life physician-patient conversations in a prospective trial.^17,18^ Considering these results, we hypothesized that AI models could predict radiation oncology patient 30-day mortality using EHR data, and that models trained with curated, comprehensive features would outperform models trained only with patients’ ages and physicians’ assessments of their overall health as documented by their performance statuses.

## Methods

Clinical features known or believed by a radiation oncologist to be associated with mortality risk were abstracted from an EHR (Epic Systems, Verona, WI) for patients who consulted with a radiation oncologist at any of seven BJC Healthcare Siteman Cancer Center sites between June 2018 and February 2024. This study’s intent was to develop one or more machine learning (ML) model(s) that may provide actionable, interpretable outputs at the time of RT consultation; therefore, features were represented in ML training data at time points nearest to the RT consultation date. For example, patient age was represented as age on the date of RT consultation. For patients with multiple RT consultations, the date of the most recent re-consultation was used as the benchmark. Recognizing that some information relevant to a radiation oncologist’s management decision may not be available on the consultation date, a two-week post-consultation buffer was coded into the preparation pipeline. For example, laboratory data obtained within two weeks of consultation date were included.

### Event

The classification event was death within 30 days of a patient’s most recent RT consultation. Patient vitality status was derived from the EHR.

### Features

Features related to patient demographics, cancer and comorbid diagnoses, American Joint Committee on Cancer (AJCC) stages and treatment histories, laboratory results, medications, substance use histories, hospitalizations, performance status assessments, clinical trial participation, oncologist notes, radiologist reports, and pathologist narratives were abstracted and prepared for ML.

#### Medications

Virtually all medication prescriptions consisted of one of the top quartile most-prescribed formulations. A radiation oncologist manually reviewed these formulations and matched those deemed potentially relevant to mortality risk to one of 984 generic medication names. For example, “PEMBROLIZUMAB IVPD IN 100 ML” (the most common pembrolizumab formulation), “KEYTRUDA IV” (a brand name), and “INV-WUSM_BJH (160621) PEMBROLIZUMAB IVBP in 100 ML” (a clinical trial formulation) were matched to the generic “pembrolizumab.” Text matching was executed with a python implementation^19^ of the Levenshtein distance,^20^ and a 100% partial ratio threshold was required for a match. Matching prioritized the most relevant formulation constituents. For example, “MEROPENEM 1 GRAM/50 ML IN 0.9% SODIUM CHLORIDE” matched “meropenem” rather than “saline.” For formulations with multiple relevant constituents, the first constituent was arbitrarily matched. For example, “BICTARVY” matched “bictegravir” rather than “emtricitabine” or “tenofovir alafenamide.” Medications included systemic therapies prescribed for cancer treatments. Medications were encoded as a binary feature (prescribed or not prescribed) because of the complexity of mapping medication dosage units (e.g. milligram, capsule, “puff”) to a single representation for each medication.

#### Prior Radiation Therapy

For patients who had previously received RT, cumulative doses per irradiated anatomic region were extracted. A radiation oncologist tokenized RT course names to 2,270 unique word fragments, reviewed them, and programmatically matched fragments that identified RT target anatomy to standardized names describing anatomic regions. For example, the fragment “BRS” parsed from “C1_RT_BRS_2023” matched “breast,” the fragment “INGUINAL” parsed from “C1 L INGUINAL 20” matched “groin,” and the fragment “GK” (standing for Gamma Knife) parsed from “C2 GK 2023” matched “brain.” Past radiation doses were summed per anatomic region per patient.

#### Laboratory Tests

A radiation oncologist reviewed all uniquely protocoled laboratory tests, combined tests that evaluated common biologic parameters, and selected tests known or believed to be associated with mortality. For example, “GLUCOSE” and “GLUCOSE BY MONITOR DEVICE POC BJH” (a point-of-care glucose fingerstick) test results were combined under “GLUCOSE,” while “NUCHAL TRANSLUCENCY” (a fetal ultrasound parameter deemed unlikely to be associated with 30-day mortality) was excluded from the ML training/testing datasets.

#### Performance Statuses

Physician assessments of Karnofsky Performance Status (KPS) were extracted for every patient. Because KPS is not a structured data element in our institution’s EHR, we wrote regular expressions to recognize templated language in radiation oncologists’ consultation notes and extract KPS scores. For some patients, the consulting radiation oncologist recorded an Eastern Cooperative Oncology Group (ECOG) performance status rather than a KPS, in which cases the ECOG score was converted to an approximate KPS. A minority of patient radiotherapy consultation notes did not document a performance status assessment. For these patients, the most recent KPS recorded by an interdisciplinary oncology provider’s note (e.g. medical or surgical oncology) was extracted.

#### Substance Use Histories

Patients’ tobacco, alcohol, and illicit drug use were extracted in structured classifications. These were reviewed and aggregated for simplicity to denote current, former, or no use.

#### Cancer Stages

The AJCC tumor (T), nodal (N), and metastatic (M) stage descriptors vary between cancer histologies and between AJCC staging versions within a single histology. To reduce staging complexity, histology-specific TNM descriptors were subsumed in a simpler descriptor subset that could more homogenously describe all cancer types. For example, T1, T1a, T1b, etc. were mapped to T1. Clinical (c) and pathologic (p) cancer stages were coded distinctly (e.g. cN1 and pN1) to recognize precise prognostic information available in post-surgical, pathologic staging while preserving clinical staging information for patients who never achieved a surgical treatment.

### Machine Learning

Missing data were imputed by means of quantitative features with a nearly normal distribution (e.g. albumin), by medians of features with a skewed distribution (e.g. creatinine), or by modes of categorical features (e.g. ethnicity). However, features for which missing data communicated clinical meaning were not imputed by a statistical measure of centrality; for example, missing breast RT doses were imputed by zeros, because patients with missing values for this feature had not received RT targeted to the breast.

ML training and testing was conducted using the python Scikit-Learn package,^21^ version 1.4.2. The methodology is visualized in Figure 1a. Data were split 70/30 into training and test datasets. Features were normalized, and the feature space reduced to 1,000 features with the highest ANOVA F variance ratios. Using the training dataset, a five-fold cross-validation grid search iteratively trained logistic regression (LR), random forest (RF), and gradient boosting decision tree (GBD) classifiers with varying hyperparameters to identify hyperparameter combinations that optimized model performance for each algorithm. Optimized models were then presented with the test dataset, and their performances quantified by accuracy, precision, recall, and F1 scores. Separately, models trained only with KPS – representing physicians’ global assessments of imminent mortality likelihood – and age were tested and compared to the comprehensive models.

**Figure 1:**
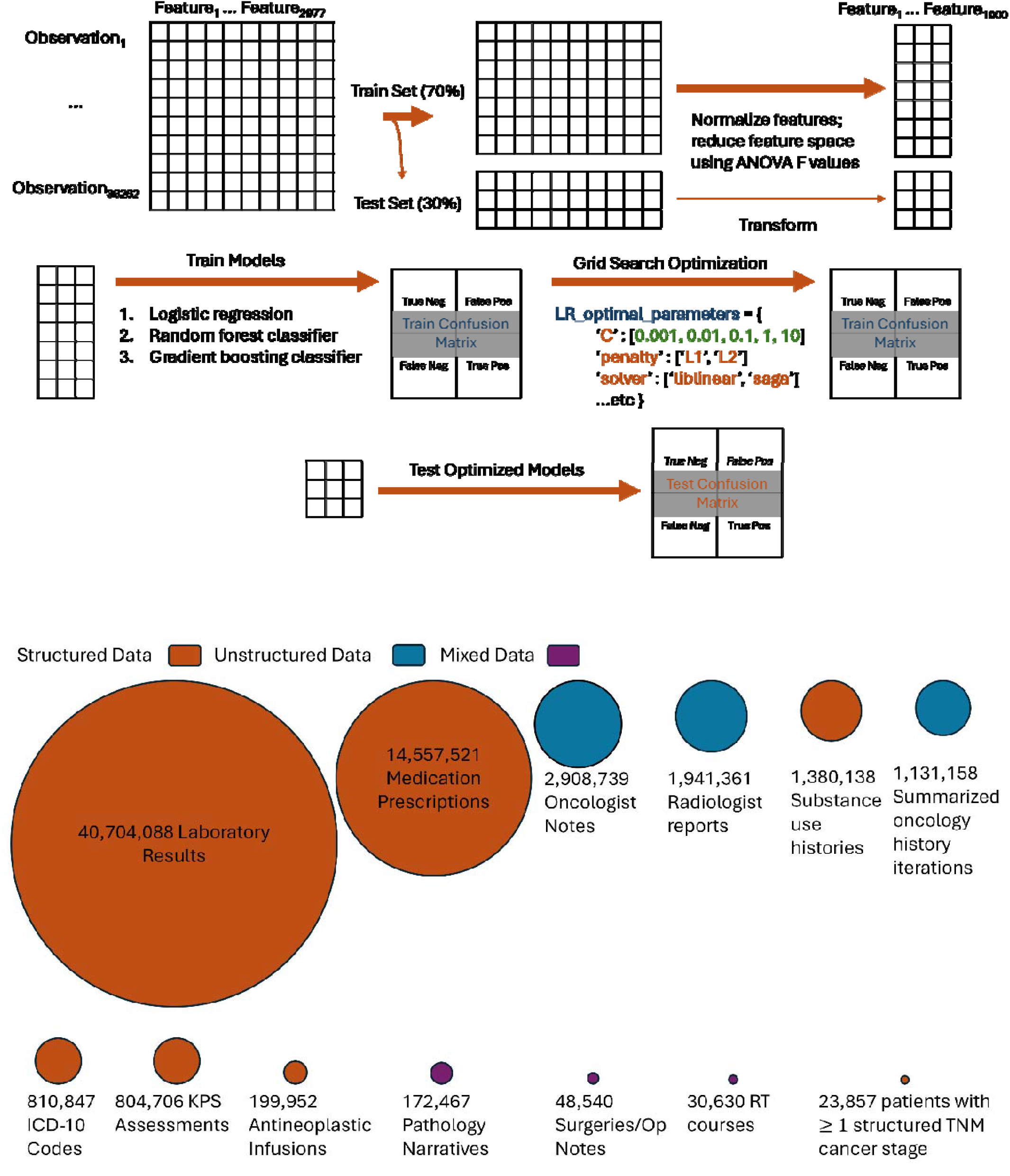
Machine learning training and testing methodology (A), and summary of abstracted EHR data from which features were selected or derived (B). ICD-10: International Classification of Diseases-10; KPS: Karnofsky Performance Status; Op: operative; RT: radiation therapy; TNM: tumor, node, and metastasis.

## Results

Clinical endpoints visualized in Figure 1b were abstracted for 38,262 patients. These comprised 34.5 Gb of tabular data, including approximately 5.3 million patient encounters, 40.7 million laboratory values resulted from any of 4,134 laboratory tests, 14.6 million medication prescriptions from any of 24,080 uniquely named medications formulations (of which the top quartile accounted for 99.2% of prescriptions), 2.9 million oncologist notes, 1.9 million radiologist reports, 810,000 International Classification of Diseases-10 (ICD-10) comorbid diagnoses, 170,000 pathology narratives, 49,000 operative notes, and 31,000 RT courses (for which 99% of treatment targets could be described by one of 45 anatomic regions).

A subset of patient characteristics is detailed in Table 1. The median patient age was 66 years, most of whom were female (52%) and white (82%). The great majority of patients had a good or excellent KPS (three-quarters scored 80 or better). Most had not previously been a hospital inpatient at the time of RT consultation, and only 12% had been hospitalized within the preceding 30 days. Few were annotated as currently smoking (12%), using alcohol (32%) or using illicit drugs (14%). Few were enrolled in an oncology clinical trial (5%). Most patients (62%) had one or more TNM cancer stage(s) that had been documented in a structured format by an oncology provider. The number of patients who died within 30 days of RT consultation was 951 (2.5%).

**Table 1:**
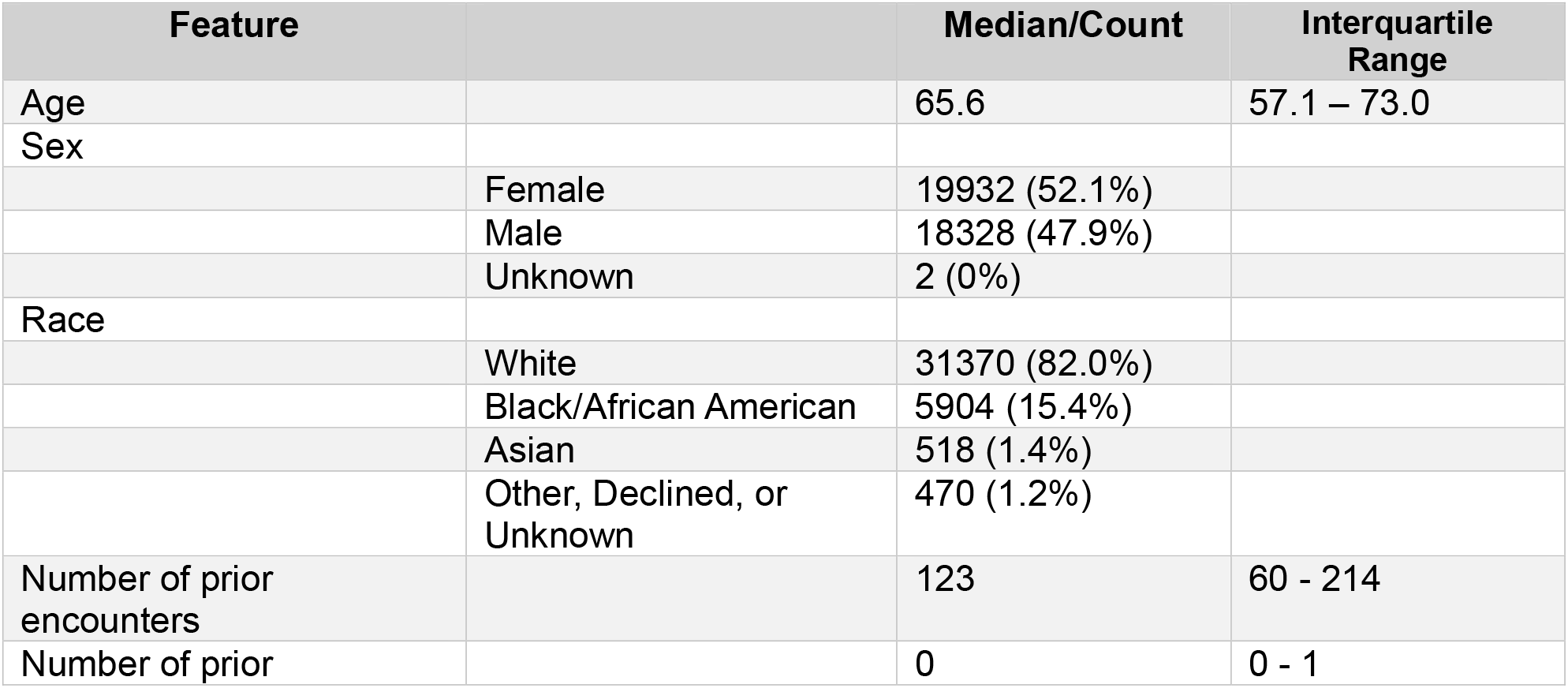
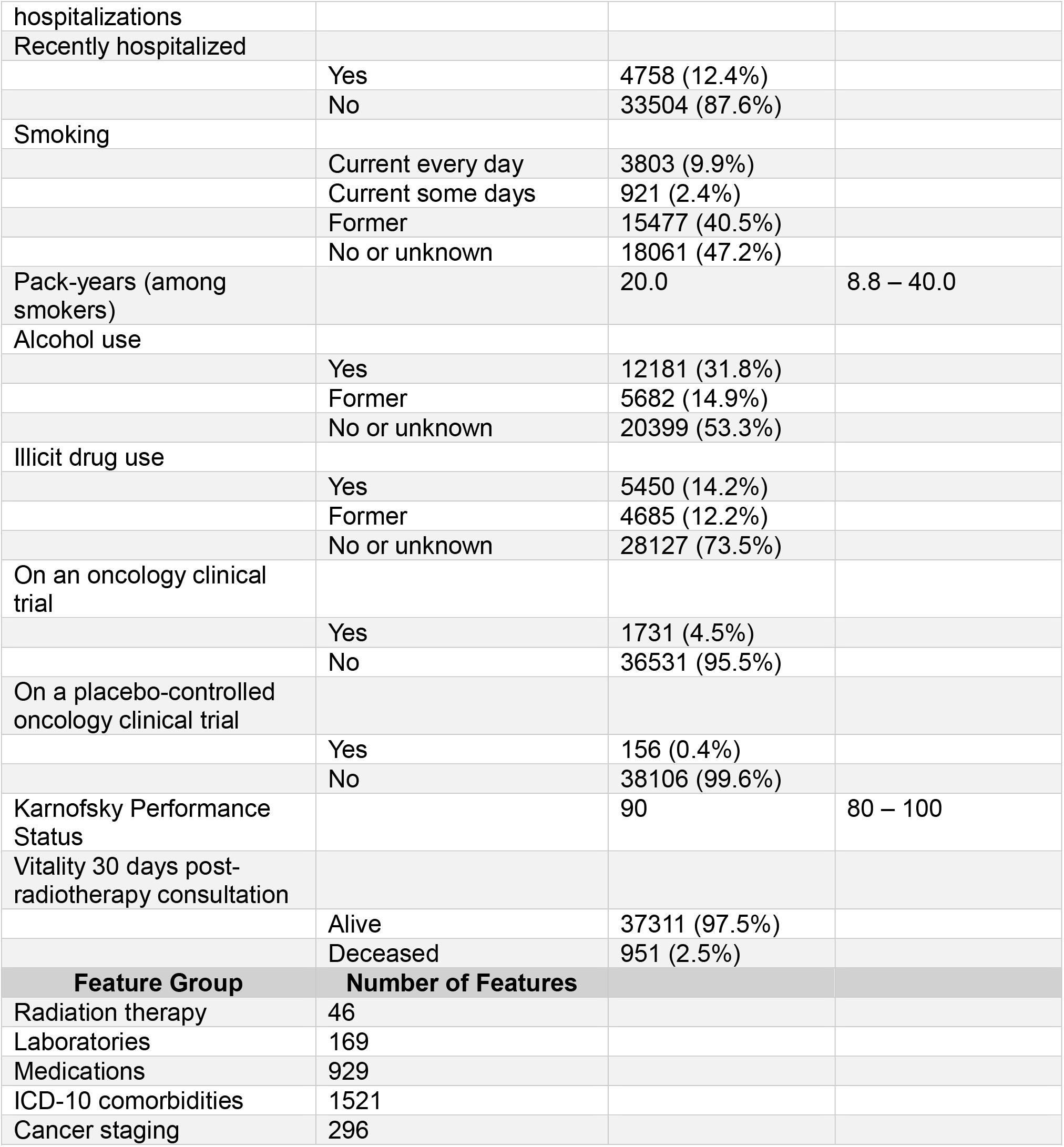
Patient cohort characteristics and selected features.

After feature selection, engineering, and cleaning, the preliminary ML dataset comprised 38,262 patients as rows and 2,977 features as columns. As described in the methods, this dimensionality was subsequently reduced to 1,000 features for training and testing.

Using an event probability classification threshold of ≥ 0.2, LR, RF, and GBD models each tested with excellent accuracy (≥ 97%). The RF model tested with the highest precision, or positive predictive value (0.53), while the LR model tested with the highest recall, or sensitivity (0.58). The RF model achieved the best harmonic mean of precision and recall (F1 score of 0.54). Model performances are detailed in Table 2.

**Table 2:**
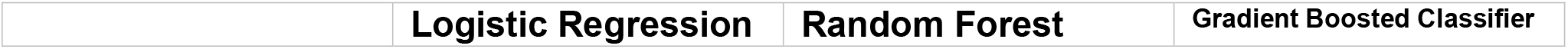
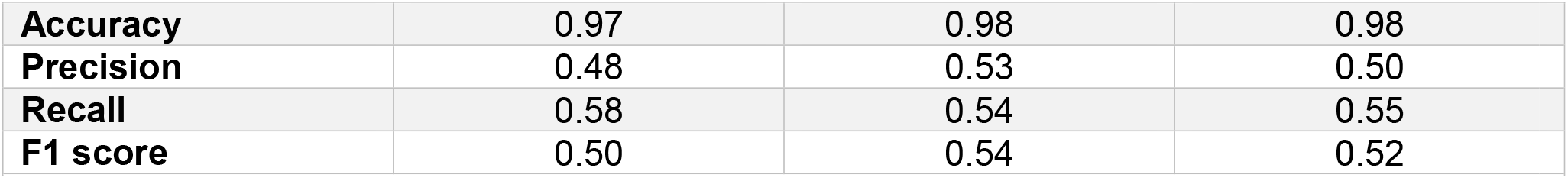
Performance of three ML models trained with 1000 clinical features to predict patient mortality within 30 days of RT consultation. Model output event probabilities of ≥ 0.2 were classified as predicted events. Accuracy is the sum of true positive (TP) and true negative (TN) predictions divided by the sum of TP, TN, false positive (FP) and false negative (FN) predictions. Precision is the TP divided by the sum of TP and FP. Recall is the TP divided by the sum of TP and FN. The F1 score is twice the product of precision and recall divided by the sum of precision and recall.

|  |  |  |  |
| --- | --- | --- | --- |
|  | **Logistic Regression** | **Random Forest** | **Gradient Boosted Classifier** |
| **Accuracy** | 0.97 | 0.98 | 0.98 |
| **Precision** | 0.48 | 0.53 | 0.50 |
| **Recall** | 0.58 | 0.54 | 0.55 |
| **F1 score** | 0.50 | 0.54 | 0.52 |

The receiver operating curve for the RF model at varying probability classification thresholds achieved an area under the curve (AUC) of 0.94, and the precision-recall curve for the same model achieved an AUC of 0.55 (Figures 2a and 2c). By comparison, an RF model trained only with KPS and age achieved a receiver operating AUC of 0.61 and a precision-recall AUC of 0.06 (Figures 2b and 2d).

**Figure 2:**
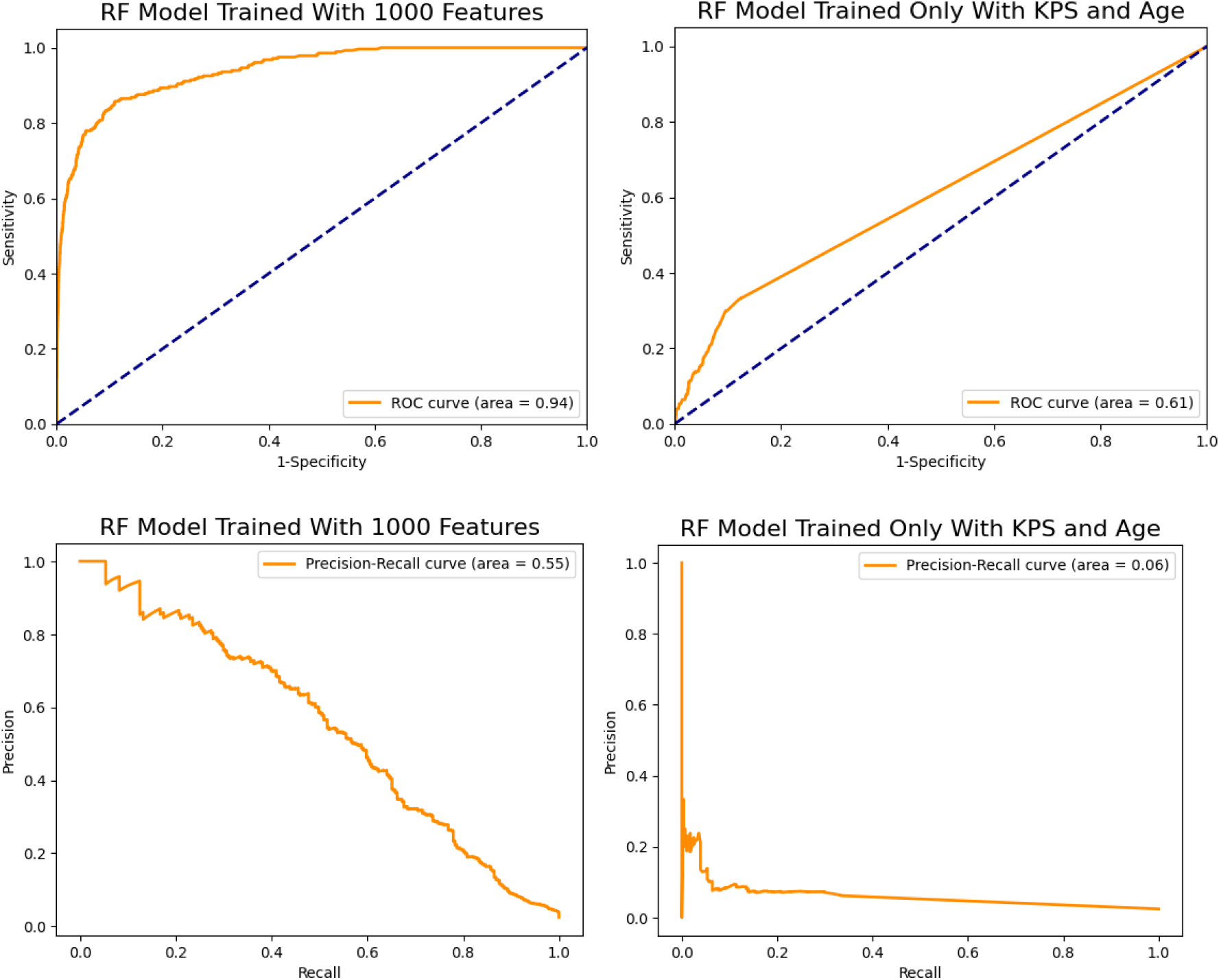
Receiver operating (2a, 2b) and precision-recall (2c, 2d) curves for random forest models trained with a full complement of 1000 curated features (2a, 2c) or trained only with patient Karnofsky performance status and age (2b, 2d).

The ten features most strongly correlated with 30-day mortality for RF, GBD, and LR models are reported in Table 3. Each algorithm independently identified the number of prior encounters as a feature strongly associated with 30-day mortality; a greater number was associated with survival. Laboratory values associated with liver (albumin, prothrombin time, international normalized ratio, aspartate transaminase) or kidney (blood urea nitrogen) failure and medications associated with end-of-life comfort care (hyoscyamine, glycopyrrolate) were also among the top performing features. Patient KPS and age were also among the top features in the LR model.

**Table 3:**
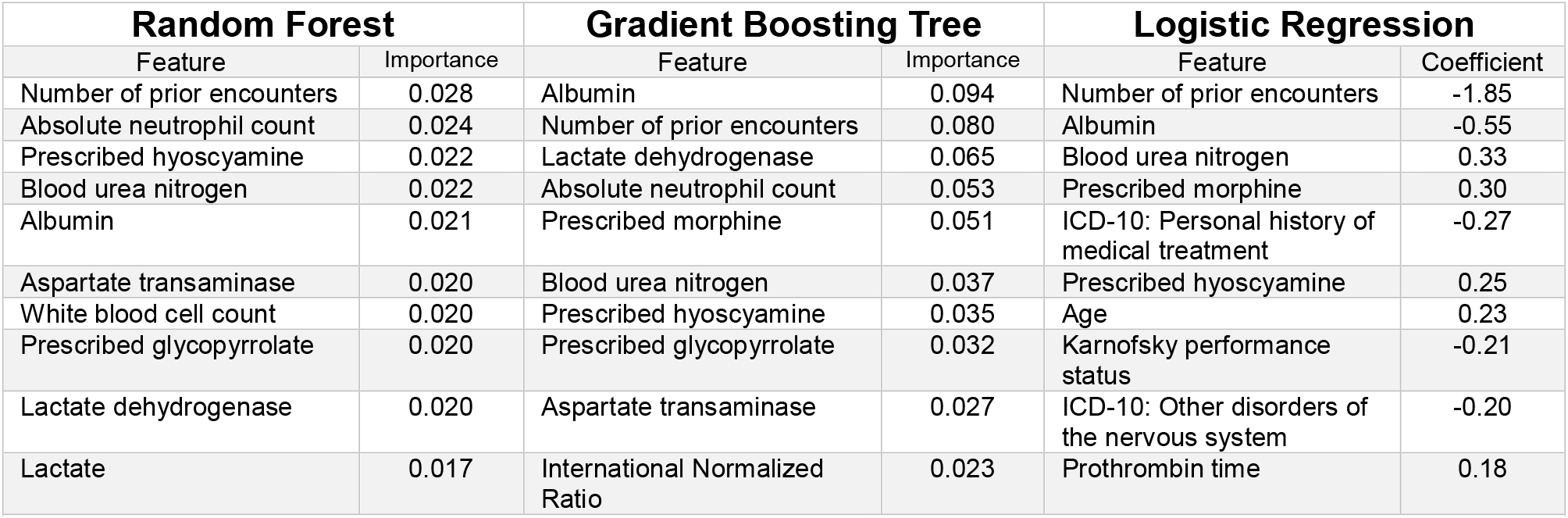
The top ten features most strongly associated with patient mortality within 30 days of radiotherapy consultation for ML models. Negative logistic regression coefficients are correlated with 30-day survival, while positive coefficients are correlated with 30-day mortality. Random forest and gradient boosting tree feature importances do not have directionality.

| Random Forest |  | Gradient Boosting Tree |  | Logistic Regression |  |
| --- | --- | --- | --- | --- | --- |
| Feature | Importance | Feature | Importance | Feature | Coefficient |
| Number of prior encounters | 0.028 | Albumin | 0.094 | Number of prior encounters | -1.85 |
| Absolute neutrophil count | 0.024 | Number of prior encounters | 0.080 | Albumin | -0.55 |
| Prescribed hyoscyamine | 0.022 | Lactate dehydrogenase | 0.065 | Blood urea nitrogen | 0.33 |
| Blood urea nitrogen | 0.022 | Absolute neutrophil count | 0.053 | Prescribed morphine | 0.30 |
| Albumin | 0.021 | Prescribed morphine | 0.051 | ICD-10: Personal history of medical treatment | -0.27 |
| Aspartate transaminase | 0.020 | Blood urea nitrogen | 0.037 | Prescribed hyoscyamine | 0.25 |
| White blood cell count | 0.020 | Prescribed hyoscyamine | 0.035 | Age | 0.23 |
| Prescribed glycopyrrolate | 0.020 | Prescribed glycopyrrolate | 0.032 | Karnofsky performance status | -0.21 |
| Lactate dehydrogenase | 0.020 | Aspartate transaminase | 0.027 | ICD-10: Other disorders of the nervous system | -0.20 |
| Lactate | 0.017 | International Normalized Ratio | 0.023 | Prothrombin time | 0.18 |

## Discussion

We demonstrated that AI models trained with ML algorithms on carefully selected, structured EHR clinical data could identify patients at elevated risk of death within 30 days of RT consultation. LR, RF, and GBD models each tested with excellent performance, but within the scope of our hyperparameter optimization methodology the RF model output best balanced precision and recall (F1 score of 0.54) at an event probability classification threshold of ≥ 0.2. It predicted with excellent AUCs for sensitivity/1-specificity and precision/recall curves (0.94 and 0.55, respectively). Moreover, we demonstrated that a physician-curated, comprehensive feature space trained better models than a space inclusive only of patient ages and oncology provider estimations of patients’ imminent mortality risk as proxied by KPS scores. The most influential features in the models’ predictions have interpretable and clinically rational mortality associations.

For context, several excellent studies have employed ML techniques to predict cancer patient mortality at various time points: a random forest model trained with 566 clinical features derived from three classes (demographics, comorbidities, and laboratories) abstracted from hematology/oncology encounters for 26,525 patients predicted death within six months of outpatient encounters with a precision of 0.513 and a receiver-operating area under the curve of 0.88.^16^ A Bayes point machine trained with 16 clinical features from 1,790 patients with spinal metastatic disease predicted 30-day mortality with a sensitivity/1-specificity AUC of 0.78.^22^ A regression model trained with “routine” laboratory, demographic, and biometric hospital admission features prospectively predicted deaths within 45 days of hospital admission for 911 solid tumor patients significantly better than the patients’ admitting providers (AUC 0.83 vs. 0.75, p < 0.001).^23^ A Cox proportional hazards model trained to predict survival time with 3,813 features derived from laboratories, vitals, ICD-9 codes, current procedural terminology codes, medications, and clinical notes (represented as 1-2 word phrases) for 14,600 metastatic cancer patients predicted one-year mortality with a sensitivity/1-specificity AUC of 0.77.^24,25^

Our study is strengthened by its feature engineering methodology, which leveraged clinical domain expertise to preserve information within data.^26^ Utilizing domain experts’ knowledge is more effective than relying on algorithms to learn knowledge inductively in ML classification studies,^27^ and classifiers built with end-user insight may also be better clinically received. A radiation oncologist engaged at every step of data abstraction, feature selection, feature derivation, missing data imputation, and model output interpretation. The resulting feature space is, to our knowledge, among the most comprehensive of any that has been published to predict cancer patient short-term mortality.

Many potential uses of this model may be conceived, but its development was specifically motivated by the clinical need to identify cancer patients who are best served by single-fraction palliative RT rather than prolonged or dose-intensified palliative RT. Results from the international phase III SCORAD trial are a case study illustrating this need. SCORAD investigated noninferiority of 8 Gy in a single fraction compared to 20 Gy in five fractions with respect to a primary endpoint of eight-week ambulatory rate in patients with spinal cord compression who were not candidates for surgical decompression. SCORAD stipulated that enrolled patients have a physician-assessed anticipated survival of at least eight weeks as an eligibility criterion. Despite this, (37%) patients died before eight weeks and could not be evaluated for the trial’s endpoint. The trial’s data monitoring committee increased the trial’s sample size, yet the trial narrowly failed to meet its noninferiority endpoint.^28^ Better patient mortality predictions might have altered SCORAD’s outcome. For enrollment to similar trials, an AI model such as ours may assess patient trial eligibility with greater accuracy and precision than a physician.

Our work has notable limitations. Vitality status was coded as it existed in the EHR, and patient deaths may have been underreported. The 30-day mortality rate in our patient population was lower (2.5%) than has been reported in advanced cancer patients (16%), although this difference is at least in part because our study included patients at all stages of cancer diagnoses. We attempted multiple means to obtain access to the Social Security Death Index to corroborate our death data, but unsuccessfully.^29^ There are other limitations to EHR data, such as that smoking, alcohol, and drug use are neither consistently recorded by medical staff nor reliably reported by patients. Furthermore, KPS assessments, when not recorded in radiotherapy consultation notes, were extracted from an oncology note that antedated the radiotherapy consult and consequently may not have represented the patient’s performance status on the date of radiotherapy consult.

There remain opportunities to improve our feature space. Only a small fraction of unstructured data was structured at scale and included in the feature space. Large language AI models (LLMs) might facilitate greater use of these data. Dagdelen et al. used GPT-3 (OpenAI, San Francisco, CA) and Llama-2 (Meta, Menlo Park, CA) LLMs to extract complex material properties as structured outputs from scientific texts for downstream ML applications.^30^ In future work, we intend to investigate whether LLM-extracted structured features — particularly related to clinically validated prognostic indices — improve our model’s performance and increase its credibility to physician end-users. For example, LLMs might extract the number of brain metastases from magnetic resonance imaging radiologist reports, which is a parameter established in the Graded Prognostic Assessment tool^31^ to be associated with survival in many cancer histologies.

Finally, we aim to test our model in a prospective clinical trial to evaluate its influence on radiation oncologist decision-making and patient quality-of-life. A May 2024 systematic review identified only 86 publications detailing randomized controlled trials evaluating AI tools in clinical practice (the review included trials published between January 2018 and November 2023). Of these, only three trials were targeted to oncology populations^18,32,33^ and only one^18^ in a palliative clinical context. Clinical validation of AI tools in oncology clinical practice broadly and radiation oncology specifically remains a critical need.

## Conclusion

An RF model trained with structured EHR clinical features from a population of radiotherapy patients at a single large academic institution identified patients at elevated mortality risk within 30 days of radiotherapy consultation with excellent accuracy and precision, and better than a model that relied only on patient age and a physician’s estimates of the patient’s performance status. This model may identify patients for whom single-fraction palliative RT courses would be preferred to prolonged RT courses.

## Data Availability

This project involved secondary analysis of electronic health records. No data were collected as primary research data. To maintain patient record confidentiality, no data will be made publicly available.

